# Access to sexual and reproductive services: Experiences of Venezuelan Migrants in Ecuador

**DOI:** 10.1101/2025.09.18.25336076

**Authors:** José Andrés Ocaña Navas, Susana Guijarro, Gabriela León, Ana Lucía Torres, Patricio Murgueytio, Odalys Heredia-Andino, Paula Yánez-Sánchez, Sarah Iribarren

## Abstract

Until 2023 approximately 7.2 million Venezuelans left their country due to political and economic crises. Ecuador was the third-largest recipient of Venezuelan migrants in the context of this ongoing human mobility crisis. Most people in this scenario experienced many vulnerabilities related to health services access, specifically Sexual and Reproductive Health, this represents risks on their health. The objective of this study was to understand the use of SRH services at the level of perceptions of access through the migratory journey; usage patterns and characteristics of SRH services; experiences of discrimination and mistreatment, and recommendations for improving access, aiming to improve the accessibility and utilization of SRH services by various populations experiencing situations of human mobility. To achieve this, we conducted focus groups with semi-structured interview guides with women, men, and people and those who self-identified as belonging to the LGBTQ+ community. We found that most participants encountered barriers to access SRH services throughout their migratory journey, while on Ecuador, men reported not using much SRH services, on the other hand women and teenage solicited mainly obstetric and contraceptive needs and the LGBTIQ+ community said that the services couldn’t meet their needs. The three groups of participants reported experiencing discrimination because of their nationality, however, women that had obstetric attention reported mistreatments because of bad medical practices that was directed to Ecuadorian women as well and the LGBTIQ+ reported being targeted mainly for their sexual/gender identity rather than their nationality. This study encompasses experiences along the migration route and in the destination country, helping to identify the underlying structural and contextual factors of these issues and suggests potential future actions to ensure unrestricted sexual and reproductive health rights.

## INTRODUCTION

In recent years, a significant portion of the Venezuelan population has fled their homeland due to ongoing severe political and economic crises. This has led to what some describe as a massive “exodus,” with approximately 7.2 million Venezuelans (approximately 20% of the total population) having left their country as of 2023 [1,2]. The majority, approximately 6 million, have sought refuge in various Latin America countries. According to the United Nations High Commissioner for Refugees Ecuador has received the third-largest number of Venezuelan migrants, approximately 513,000, during this ongoing human mobility crisis [3]. In mid-2021, during the height of the COVID-19 pandemic, Ecuador was home to approximately 430,000 individuals of Venezuelan nationality [4]. Among these migrants, nearly half were women (46%), mostly of reproductive age (between 18 and 45 years old) (92%), and less than half (43%) had completed a secondary education [5]. The most recent migration wave to Ecuador primarily included individuals with more extreme vulnerabilities, including economic hardship, health concerns, limited education, and lack of access to adequate housing or arriving with no housing options [4–6].

Amid these challenging social conditions, the ongoing pandemic exacerbated the vulnerabilities faced by Venezuelan women (girls, adolescents and adults) in a situation of human mobility causing them to be particularly vulnerable to unintended pregnancies, gender-based and sexual violence, as well as sexually transmitted diseases, (STDs) and HIV/AIDS [5,7,8]. Although there are less evidence available, adolescent and adult men also faced conditions of vulnerability in this context [5,9]. Studies conducted among Venezuelan migrant women on the border with Colombia found that these women were disproportionately subjected to various forms of gender-based and sexual violence, making them more vulnerable than men in situations of human mobility [10]. Additionally, women often faced substantial challenges accessing health services due to legal constraints, discrimination, and a lack of adequate information[10]. Furthermore, conservative stances related to sexual and reproductive health (SRH) were considered a significant threat to the reproductive rights of these women [10]. On the border with Brazil, substantial barriers were identified in accessing long-acting contraceptive methods, hindering the exercise of their reproductive rights [11]. Currently, Ecuador lacks comprehensive information on the accessibility of SRH services for individuals in situations of human mobility. To address this gap, we conducted a study to explore the challenges faced by Venezuelan migrants in a situation of human mobility residing in the city of Quito. The primary objective was to identify strategies to improve the accessibility and utilization of SRH services among diverse migrant populations. This manuscript presents the qualitative findings related to SRH service use, with a focus on a) perceptions of access along the migratory journey; b) patterns and characteristics of SRH service utilization; c) experiences of discrimination and mistreatment, and d) participant-driven recommendations for improving access.

## METHODS

### Study overview

This qualitative descriptive study, using focus group and in-depth individual interviews, was part of a larger mixed methods study conducted from September 2020 to May 2022. The study focused on understanding the experiences of individuals from Venezuelan in a situation of human mobility residing in Quito. The investigation was guided by the Immigrant Health Service Utilization (IHSU) framework [12].

### Study team

The first author, Dr Ocaña, a male researcher with specific expertise in qualitative research and mixed methods and is affiliated with civil society organizations that support migrant populations. They received support from Dr. Iribarren, a female U.S. nursing scientist with approximately 16 years of research experience in Latin America (Argentina, Ecuador, and the Dominican Republic), who has mentored early-career researchers and supported research teams employing mixed methods. The overall team was supervised by Dr. Guijarro, a female physician specializing in adolescent health with more than 30 years of experience in SRH services in Ecuador. She served as the lead Principal Investigator. Dr. Murgueytio, a male physician specialized in public health with extensive experience in health services administration.

The entire process was gender-sensitive and accounted for human mobility phenomena, guided by Dr. Torres, a female sociologist specializing in gender studies. Yánez and Heredia are early-career female researchers actively involved in managing this study.

Throughout the data collection, analysis, and writing process, the research team engaged in reflective practices as part of the analytical approach. No individuals other than the researchers and participants were involved in any aspect of this study.

### Participant recruitment

Purposeful and snowball sampling techniques were used. The study included adolescents and adults over the age of 14 who were of Venezuelan nationality, self-identified as migrants, had resided in Ecuador for at least one year, and had accessed sexual and reproductive health services for any reason. Participants were contacted in three locations.

Participant recruitment commenced on October 25, 2021, and concluded on March 7, 2022. Focus group sessions were conducted within a fifteen-day window following recruitment phases, initiating on November 17, 2021, and concluding on March 19, 2022. Data collection was carried out across three distinct geographical sectors within Quito: the southern zone (Chillogallo neighborhood), the central zone (Isidro Ayora Gyneco-Obstetric Hospital), and the northern zone at the facilities of a non-governmental organization (NGO) providing support to the Venezuelan migrant population. Written informed consent was obtained from all participating adults. For adolescents under 18 years of age, informed assent was secured from the minors alongside written consent from their legal guardians.

Researchers contacted and invited participants both in person and by phone at the three locations, with the support of healthcare professionals from the health centers and the NGO. In some cases, these participants facilitated the recruitment of other eligible community members by informing them about the study and encouraging them to meet with the researchers and participate.

### Data collection

From October 2021 to March 2022, we conducted nine focus group discussions (FGDs) with the following distribution: two groups of heterosexual adult and adolescent women, two groups of heterosexual adult and adolescent men from community settings, two groups of adult women who were users of health services, one group of adolescent female health service users, and two groups of LGBTQ+ individuals.

Prior to participation, informed consent was obtained from all participants. Each FGD began with an overview of the study objectives, an opportunity for participants to ask questions, and an introduction to the interviewers. The research team developed semi-structured interview guides, which were adapted to address the specific characteristics of each group. The guides covered key topics, including perceptions of SRH status throughout the migration journey, access to SRH services, experiences with SRH care and service utilization, and participants’ recommendations for improving SRH access. As the discussions progressed, new themes emerged and were incorporated into the study findings.

All study procedures were conducted in Spanish. Each session was audio-recorded, and field notes were taken by the research team. FGDs lasted approximately 120 minutes. Data collection, including interviews and focus groups, was concluded upon reaching data saturation.

Data collection through interviews and focus groups consisted of a single session per participant or group, with no repeated measures or follow-up sessions conducted.

### Theoretical Framework that guided thematic analysis

We used the Health Service Utilization (HSU) framework proposed by Yan and Hwang [13] to analyze the utilization of SRH services by Venezuelan migrants. This framework considers factors such as health care needs, available resources, predisposing factors, and macrostructural and contextual conditions. It also helps elucidate the mediating relationships among these factors, both in general and within the specific context of migrants [13].

Additionally, we incorporated insights from Regts’ study [14], which examined health-seeking behavior through four categories: *cumulative exposure to mistreatment* (measured using a scale that quantifies the collective amount of healthcare mistreatment experiences reported by the participant), *discrimination-based attributions for mistreatment* (how participants attribute mistreatment to discrimination), *negative emotions* (the degree to which participants experience negative emotions due to mistreatment), and *continuity of care* (ongoing and consistent healthcare access). Sheferaw et al., [15] further categorized mistreatment of women during childbirth into four domains: physical abuse, verbal abuse, failure to meet standards of care, and poor rapport between women and providers. These categories aid in identifying attitudes in health service usage that can be classified as abusive or indicative of deficient health care.

D’Oliveira et al., [16] explain violence against women in health care institutions through the following categories: neglect, verbal violence, physical violence, sexual violence, professional context (including the socialization of health-care professionals and broader societal issues affecting health care delivery), and prevention interventions (actions in recruitment, training, socialization processes in professions, and improvements in working conditions). These theoretical perspectives provided a comprehensive framework for our thematic analysis enabling us to identify and categorize various forms of mistreatment and deficiencies in SRH usage.

### Analysis

The qualitative data were analyzed utilizing a thematic content analysis approach, facilitating the identification of salient patterns and thematic constructs embedded within participant narratives.

The audio-recordings were transcribed verbatim and then independently coded both inductively and deductively [17] by two research team members with qualitative coding expertise (JAO, GL). The data was coded using Atlas Ti (version 22), a qualitative data processing software, with continual refinement during the coding process to help elicit and identify themes. Initially, the researchers read through the interviews independently, performing open coding to capture initial themes and insights. They then moved to axial coding, refining and relating the codes to each other to identify central themes. After the identified codes and themes were organized and mapped onto the theorical categories described above, we added emergent categories, and we structured the findings in alignment with the analytical framework. Two additional authors (SG and SI) reviewed the initial codes and themes to ensure consistency and comprehensiveness. The research team held weekly meetings to discuss the codes and themes, achieving consensus through collaborative review.

The results were obtained through a rigorous analysis conducted by the research team, followed by a review and validation process involving adult participants and LGBTQ+ subject matter experts. This participatory approach ensured the findings’ accuracy and faithful representation of participants’ lived experiences. The term “migratory trajectory” describes participants’ journeys from their places of origin to their arrival in Quito, encompassing the diverse risks, needs, and health conditions related to barriers in accessing SRH services.

### Ethics

The University Pontificia Universidad Católica del Ecuador (PUCE) Institutional Review Board (CEISH-878-2020, Code PV-01-2020) the PAHO Ethics Committee (PAHOERC Ref. No. PAHOERC.0330.02) approved this research for ethical content and management and the Health Intelligence Directorate of the Ministry of Public Health (MSP-SNSG-2020-15725).

## RESULTS

The study sample consisted of 61 Venezuelan migrants residing in Ecuador for an average of 3.1 years (range 0.7-4.3), ranging in age from 10 to 43 years. Of the participants, 54.1% were women and 19.7% identified as part of the LGBTIQ+ population. Across all gender groups, the majority had attained secondary education; however, the LGBTIQ+ group had lower overall educational levels. Most participants lived in southern Quito, in urban-marginal areas characterized by socioeconomic vulnerability. While most women were recruited through healthcare services, heterosexual men and individuals from the LGBTIQ+ community were primarily reached through community-based or non-healthcare-related settings. No eligible participants declined to participate or withdrew from the study.

**Table 1.**
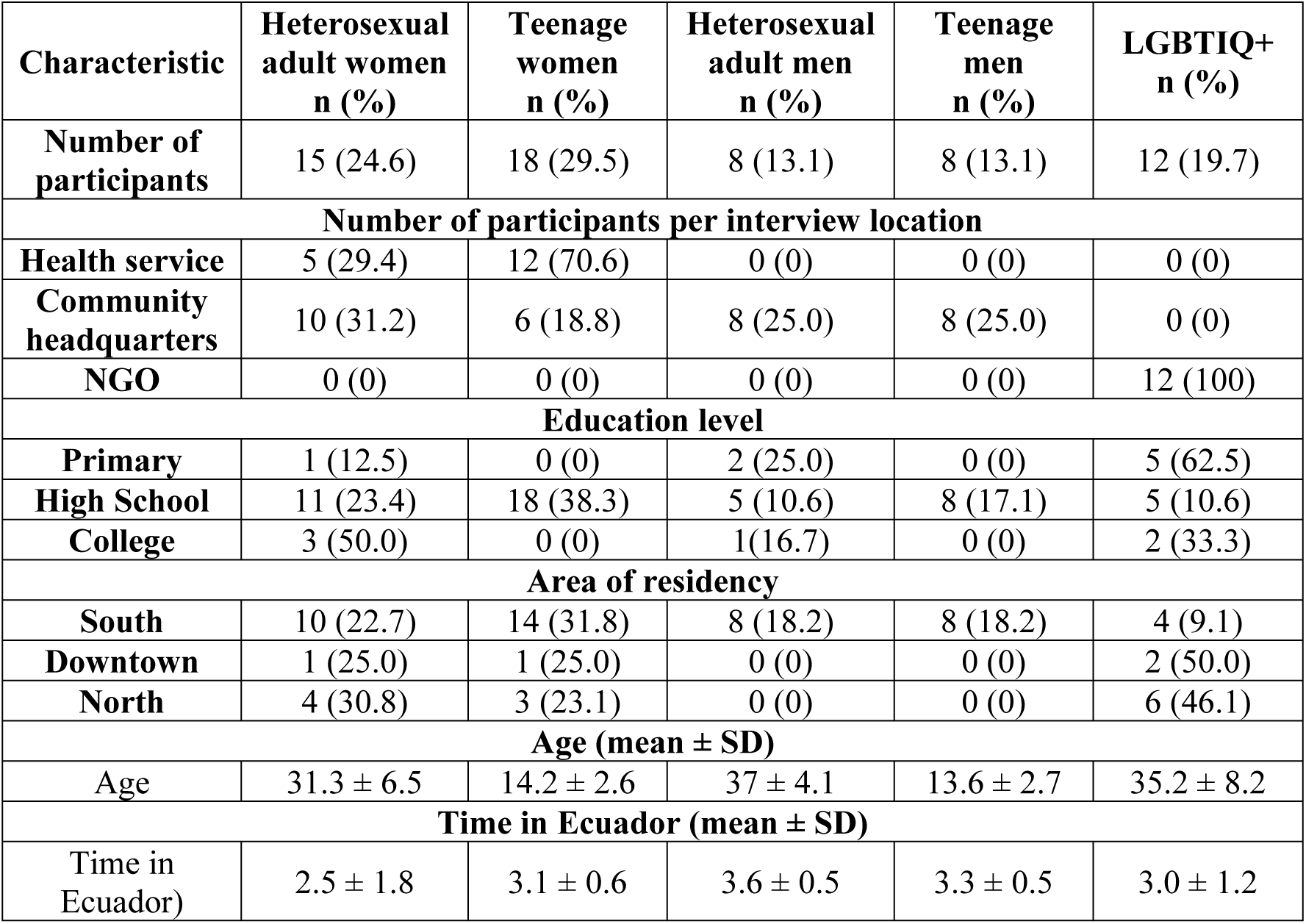
Participant characteristics (n=68)

| Characteristic | Heterosexual adult women<br>n (%) | Teenage women<br>n (%) | Heterosexual adult men<br>n (%) | Teenage men<br>n (%) | LGBTIQ+<br>n (%) |
| --- | --- | --- | --- | --- | --- |
| **Number of participants** | 15 (24.6) | 18 (29.5) | 8 (13.1) | 8 (13.1) | 12 (19.7) |
| **Number of participants per interview location** |  |  |  |  |  |
| **Health service** | 5 (29.4) | 12 (70.6) | 0 (0) | 0 (0) | 0 (0) |
| **Community headquarters** | 10 (31.2) | 6 (18.8) | 8 (25.0) | 8 (25.0) | 0 (0) |
| **NGO** | 0 (0) | 0 (0) | 0 (0) | 0 (0) | 12 (100) |
| **Education level** |  |  |  |  |  |
| **Primary** | 1 (12.5) | 0 (0) | 2 (25.0) | 0 (0) | 5 (62.5) |
| **High School** | 11 (23.4) | 18 (38.3) | 5 (10.6) | 8 (17.1) | 5 (10.6) |
| **College** | 3 (50.0) | 0 (0) | 1 (16.7) | 0 (0) | 2 (33.3) |
| **Area of residency** |  |  |  |  |  |
| **South** | 10 (22.7) | 14 (31.8) | 8 (18.2) | 8 (18.2) | 4 (9.1) |
| **Downtown** | 1 (25.0) | 1 (25.0) | 0 (0) | 0 (0) | 2 (50.0) |
| **North** | 4 (30.8) | 3 (23.1) | 0 (0) | 0 (0) | 6 (46.1) |
| **Age (mean ± SD)** |  |  |  |  |  |
| **Age** | 31.3 ± 6.5 | 14.2 ± 2.6 | 37 ± 4.1 | 13.6 ± 2.7 | 35.2 ± 8.2 |
| **Time in Ecuador (mean ± SD)** |  |  |  |  |  |
| **Time in Ecuador)** | 2.5 ± 1.8 | 3.1 ± 0.6 | 3.6 ± 0.5 | 3.3 ± 0.5 | 3.0 ± 1.2 |

Findings were categorized into three main themes: perceptions of access to SRH services along the migratory journey, characteristics of use of the SRH services in Quito, and perceptions of discrimination and mistreatment in Quito. The following describes the themes and subthemes within each category and corresponding exemplars.

### Perceptions of access to SRH services along the migratory trajectory

#### Venezuela

Participants from the groups (LGBTQ+, women, and men and adolescents of two sex) reported significant barriers to access SRH services throughout their migratory journey. In Venezuela, the health care system was perceived as being in poor condition before their departure, marked by a shortage of essential supplies, medicines, and limited access to healthcare professional care in public health services. Consequently, they were forced to seek care in the private healthcare system, incurring substantial expenses. One participant shared, “Now if a person needs an operation, you must take all of the supplies, if you need blood you have to buy the concentrate…I am being attended in a private healthcare setting [for prenatal care], but I am going to give birth in the maternity hospital [public] because I am going to have a c-section, from now on I am must buy everything, even to sew the wound, all because if I do not take everything for the delivery, they will not attend to me” (Adult female service user).

#### Colombia

In Colombia, participants encountered difficulties accessing healthcare services, primarily due to the complexities of the health insurance system. They found the system inadequate for their needs, necessitating self-care to avoid reliance on healthcare services. A participant noted, “[In Colombia] it was difficult to enter a hospital to give birth…and I couldn’t…if I missed the appointment, I had to spend three [or] four days to get another appointment” (Teenage female service user).

#### Colombian - Ecuadorian Border

As individuals progressed through their journey, and before the pandemic, at the Colombian - Ecuadorian border, they received quality support from civil society organizations, including medical care, food, and hygiene assistance. One participant shared, “When I arrived to Ipiales at the border with [Ecuador]…the Red Cross was already there providing care in a tent. They helped me taking care of my daughter excellently (…) they gave me water, (…) they were very attentive and not only with me, to everyone who was passing by they gave medical intervention” (Adult woman).

#### Ecuador

In Ecuador, access to healthcare is considered “universal,” however the participants’ experiences varied by the quality of care provided and the specific reasons for seeking care. Positive experiences included, for example, “I needed eye surgery in the middle of the pandemic, and I was able to receive it. I went to the emergency room twice for kidney stones, and I was well cared for” (Adult woman). Conversely, a negative experience was shared by another participant, “My baby was born here, he is Ecuadorian, and she [wife] was sent to do all her check-ups at the Maternity Hospital, very well there, they treated her very well. However, when it is the most basic health [primary care], we who have children, as they say, primary local healthcare is, they don’t receive you. I went and up there [ to the primary care center], I had an argument with the doorman, he said, ‘no, you can’t go in with the child (of seven months)” (Adult men).

#### Impact of the COVID-19 Pandemic

During the pandemic, various health services redirected their resources, leading to closure of SRH services, limited access to healthcare, and restricted emergency care. A teenage woman shared, “Before everything was open, the operating rooms, everything, and due to the pandemic they had to close many centers, they did not want to treat my son and me, they said ‘that’s not COVID and we are not going to give you attention here, so leave because we are not going to attend you here.”

Similarly, at the Colombian-Ecuadorian border, healthcare services were suspended, particularly for adult women and adolescents. One participant reported, “I arrived at the frontier, and it was closed. I was going down a trail and some people grabbed me, I thought I was going to die on that trail, which was the worst. We were afraid that they would do something to us, or we would get into an accident” (Teenage woman).

#### Characteristics of the use of the SRH services in the City of Quito

##### Meeting health needs

SRH care needs varied among each of the groups, highlighting the particular health needs expressed by each group.

##### Heterosexual adult men

Most heterosexual adult men expressed limited demands for general health services and SRH services, prioritizing their care as secondary. One participant shared, “…gender disparities and traditional masculinity is also a reason for not seeking care promptly. We wait until things become serious. For example, that thing about rectal examination, the prostate exam, which makes you uncomfortable, you say you don’t want to go because it’s weird, because it is sexist to not take care of yourself and have prejudices” (Adult man).

When men do visit SRH services, it is typically either as companions to their partners or for specific urological concerns, such as fertility issues, vasectomies, and STDs. They typically seek attention in specialized healthcare services rather than primary care. For example, one stated, “I went to the clinic, because in the public there were never available dates, but I went after a long time and at first I was embarrassed to take down my pants, it is difficult for another man to check you” (Adult man).

Regarding vasectomies, some participants sought this procedure but encountered challenges. They couldn’t find it within the public health system, and in the private system, the cost made it financially inaccessible. One participant explained, “I wanted to have a vasectomy, but in hospitals and health centers they don’t do it and in the clinic, I couldn’t afford it, my boss wanted to help me, but that meant deducting it from my salary, which wasn’t convenient” (Adult man).

##### Adolescent men

Adolescent men stated they rarely visit healthcare services, and when they do, it’s typically with their peers. They expressed a need for access to contraceptives and an interest in spaces for comprehensive sexual education, particularly in school environments among peers of the same age. Although this need may be addressed by SRH services in primary care and schools, discussions about sexuality are not openly discussed, and home is considered a more conducive space for these discussions. One adolescent man shared, “I have never gone to the health center on my own, but since some of my colleagues already have sexual relations, we do need access to condoms, it’s always embarrassing. Once, a friend asked us to go with him, so we went. I spoke with the doctor and well, when they gave me the box of condoms, they told me everything…what bothered me the most was the way the lady looked at my friend and me. When we turned around, I looked at the door, and I ran and left” (Teenage man). Another account mentioned, “In school they do mention sexual education to us, but sometimes it’s embarrassing to ask adults” (Teenage man).

##### Women and adolescent women

Conversely, adult women and adolescents reported specific SRH needs. These included maternal and childcare, obstetric care during pregnancy, childbirth, postpartum care, contraception, and abortion services. Female adolescents, in particular, expressed a need for comprehensive sexual education. One adolescent woman explained, “Back in [Venezuela], it was challenging to discuss sexuality. Young people have no one to talk to, and we end up getting pregnant. Like me, I am 18 years old, and I have a son, and he is 1 year old. I got pregnant at 16 and had him at 17. If I were there, I couldn’t ask and learn, so you can plan or take precautionary measures to avoid getting pregnant, but they only say that you have to be focused on studying. They were not looking for a way to teach us. Here in [Ecuador], it is more about comprehensive sexual education, but we still need to have more knowledge” (Teenage woman).

Another particular need reported by adult women was for tubal ligation. This service was suspended due to the pandemic. One woman stated, “Before the pandemic I wanted to have the ligation done…I no longer want to have children, it would be very difficult to have another one now, but with the pandemic they no longer perform the procedure to anyone, and it is very expensive outside [private healthcare setting]” (Adult woman).

##### LGBTIQ+

Among LGBTIQ+ participants, there were many unmet health needs, primarily due to their Venezuelan nationality and additionally their identity as part of the LGBTIQ+ community. This includes unmet needs in sexual and reproductive health. They report not receiving any response from either the state or private systems. One participant stated, “You see, in early 2020, before the pandemic, I was kidnapped and raped, I have a fissure in my rectum and I have been waiting for almost a year for an operation that they have not been able to do. I was tired of the pain. I went to a private hospital, a super expensive hospital. I went because I needed it and I didn’t know what to do, but they didn’t treat me there either, in the public hospitals they didn’t want to treat me, in both places, because I was gay” (Man of the LGBTIQ+ community). Another negative experience was, “I told the secretary who gives the appointments that I had a urinary tract infection, and she started like, judging me, but that’s not from now, that’s from a long time ago. She told me that it’s not a problem anyway” (Man of the LGBTIQ+ community).

The LGBTIQ+ participants also mentioned serious limitations in accessing differentiated and specialized care, especially for the trans population requiring hormonal and surgical treatment. These limitations stemmed both from a lack of adequate physical space for providing such care and from insufficient availability of appropriate medical services. One participant stated, “The service [of surgical and hormonal treatment] I from 2007 to 2017 in Spain. I came here to Ecuador, and I was trying through the public service to get an appointment with the endocrinologist, and they never gave me one, how can it be possible, that is why, I tell you that Ecuador is not yet ready for something like this” (Trans woman of the LGBTIQ+ community).

### Financial and social resources

#### Financial resources

##### Adult men

Adult men reported being the primary financial providers for their families, often prioritizing their family’s needs over their own health. They generally had limited financial resources to allocate towards health services. One participant explained, “I have other children in Venezuela, and how am I going to send them money if I have to go to a private health center? Because in the public system it is impossible. My salary paid every fifteen days, is not going to be enough for me, and if I stop working because I am unwell, it is not going to be enough.” (Adult man)

##### Adult Women

The Covid-19 pandemic adversely impacted both financial and social resources for women. Women reported having limited financial resources, often needing to pay out-of-pocket for exams and medication not covered by the public health system. They reported extensive social resources through networks of friendship with other women, which help them navigate the healthcare system. One woman shared, “Before the pandemic started, I was taking microgynon contraceptives and I decided to get the implant. The first time I went, there was no doctor, the second time I went I had to wait and wait, and they told me I missed my turn. The next time I went, I was already pregnant; I couldn’t always buy the pills” (Adult woman).

##### LGBTQ+ Community

Participants from the LGBTIQ+ community reported relying on specialized NGOs for their care, utilizing social resources such as oral transmission, social networks, and WhatsApp groups. However, they found more support from NGOs than from public health services. One participant stated, “The NGO that provide LGBTIQ+ focused care supported me throughout the pandemic, my experience, thank God, was different, they supported me with groceries and health assistance. It was easier to find out what the NGO was doing through social media. Everything was closed and nowhere wanted to attend you if it wasn’t COVID” (Man LGBTIQ+ community).

##### Social Resources Adult Women

Women reported an important social resource was oral transmission of information about the various services available to them, including NGOs and local government health services. During the pandemic, this information was disseminated through WhatsApp, allowing them to navigate through the constraints of confinement and physical distancing. One woman shared, “During the pandemic where everything was closed, thanks of the news among women, we met a human mobility person here who checks us in a local government clinic. She is a woman who gives us appointments through WhatsApp messages, she is very kind, and she performs exams, we all write to her and share her contact information with each other” (Adult woman).

##### Services Resources

Participants perceived that health services in Quito were experiencing a significant crisis in terms of available resources, exacerbated by the impact of the pandemic. These challenges were more evident in primary care services and occurred less frequently in specialized hospital services. Shortages: Participants noted shortages of HIV and contraceptive medications as well as general supplies. One woman stated, “Before the pandemic you could access health services and they gave you medicines, with the pandemic there was a lack of medicines, and they sometimes said that there were no doctors to treat you because they were treating COVID” (Adult woman). Specialized Hospital Services: In contrast, some participants had positive experiences with specialized hospital services. One woman noted, “I had all my pregnancy checkups at the maternity hospital without any problem, they were always there, it was more difficult for them to attend to you at the health services” (Adult woman).

Geographic availability: The geographical availability of public health services also influenced the use of services. The constant mobility experienced by Venezuelans limited their ability to schedule appointments and receive the necessary attention close to where they are living.

One woman shared, “I found out I was pregnant, they told me to call 171 and I called, and they gave me the appointment, it was three weeks later. And then I moved to other neighborhood and had to call again, they gave me an appointment for three weeks later, I started my checkups 6 weeks later” (Adult woman).

##### Predisposing factors

Sociocultural factors significantly influence the use of SRH services among Venezuelan migrants. For men, there was a noticeable avoidance or hesitation to access health services due to ingrained sexism and perceptions of masculinity. One participant shared, “Women are the ones who go to health centers with a doctor…men do not. Very few men say, ‘I’m going to go because I must have a problem, or I want to take care of myself’. As such, to do a checkup, no, because we are like very “macho” (manly). We don’t talk about this among friends either…we never recommend going to this or that doctor like women do” (Adult man).

For women, cultural traditions related to family care during pregnancy, childbirth, and the postpartum period play a crucial role in their health-seeking behaviors. Many women consider returning to Venezuela to receive care from their families, highlighting the importance of familial support during these stages. One woman shared her experience, stating, “I got pregnant here in Ecuador, but I wanted my mother to take care of my pregnancy, although in the end I didn’t go to my country because there you have to buy everything for the birth, if it’s a c – section even the needles. It is difficult to give birth without your family, your mother or your grandmother, but here we were able to give birth without spending anything” (Adult women user of the service).

##### Macrostructural and contextual factors

Macrostructural factors within health systems or other national systems also influence the utilization of SRH services. A primary factor is the availability of an Ecuadorian identity document. Having a document number is a specific requirement for scheduling medical appointments and receiving exams in the Ecuadorian public health system. Venezuelans without an identification document found it challenging to make appointments. One participant explained, “Ecuador enabled us to be with the Andean letter, with your Venezuelan ID…it has only 8 digits…you call to make an appointment and the first thing they ask you is your Ecuadorian ID [it has 10 digits]. If you enter your Venezuelan ID it doesn’t work, therefore it’s very difficult to make an appointment through the call center that is automatic” (Adult man).

##### Perceptions of discrimination and mistreatment in Quito

Participants consistently described experiencing mistreatment and discrimination when using public health services in Quito, creating a significant barrier to accessing SRH services. These perceptions were categorized at two levels: the Health services level (level of care and the type of professional providing the service) and individual characteristics level (factors such as place of origin, age, and sexual and gender identity). The types of mistreatments reported by participants fell into five distinct groups: physical abuse, verbal abuse, negligence, sexual violence, and poor relationships.

##### Level of care and type of health professional

Primary care: All participant groups reported barriers to accessing services in primary care, with a particular emphasis on verbal abuse. Health professionals were often perceived as imposing local norms of behavior through harsh communication. One adult woman recounted her experience, “I was referred to the health center…[The doctor told me], here in Ecuador things are not that way…And well, she mistreated me, she told me, you are this…how is it possible that you are irresponsible. [I answered], but doctor, if they do not do the [examinations] where you send me, I cannot bring back results. She was very harsh when saying things, she treated me as irresponsible…and [she was] derogatory” (Adult women).

Participants also reported a lack of information provided by health professionals, contributing to strained relationships. An LGBTQI+ individual shared their experience of feeling dehumanized, “Sometimes they think…like one is an object, like one has no emotions, one has no feelings, and I’m not just speaking for Venezuelans but in general, sometimes doctors can be so cold with a patient, so that’s like…a shock. For example, after I was raped, they treated me badly, they sent me and told me to go, do all of my exams and to this day I am still under that surveillance, and they continue to treat me like an object” (LGBTQI+ participant).

At the tertiary level of care, participants generally perceived less discrimination compared to primary care. They noted that tertiary-level facilities had better capacities to provide comprehensive and nondiscriminatory care. For example, an adult woman shared her positive experience, “I suffer from hypothyroidism and from there at the Health Center, they referred me to the hospital, where they did the ECO, at Hospital. There they did the ECO, the doctor saw me, and they did everything that was pertinent to that. (…) However, since I had hypothyroidism, I had to see a family doctor or else a gynecologist, so they referred me to the Vicentina Health Center, there is where I tell you, that I had a bad experience because the doctor was very prejudiced. (..) [she said] “here in Ecuador things are not like that (…) here you have to do this”, comparing, because I am not from here” (Adult woman).

However, participants still identified instances of mistreatment based on the type of health professional. A teenage women recounted her experience after a c-section, “They didn’t want to tell me where my son was, then they told me he is with oxygen, [they said] “stay still here, we are not going to kill your son”. After the first day of the c – section, after the first day of being born, in the morning they told me [nursing assistant], get up and make your bed, you have to make the bed, you don’t even help me. Women were just staring, and I said, “you are women too, you must have kids of your own, you have to understand a little bit more” and they just scolded me. Doctors arrived in the morning and treated us well; in the afternoon they [nurses] were angry” (Teenage woman).

##### Characteristics of population

Based on experiences of individuals interviewed, the perception of discrimination in accessing SRH services in Quito varies depending on gender identity, place of origin and age.

Gender identity: For the LGBTQI+ participants, discrimination is primarily perceived due to their gender identity rather than their place of origin. Participants in this group emphasized that health professionals often hold prejudiced assumptions about their health based on their gender identity. A trans women recounted her experience, “When you are homosexual, Trans or transvestite, doctors think you have HIV (…), I had pain in my stomach and the first thing the doctor asked me was if I had an HIV test, so I didn’t understand how that was necessary, nor how it was related. To examine me he put on double gloves, it wasn’t because I was Venezuelan, and some colleagues are not attended firsthand because they are transvestites rather than caring about where they are from.” (Trans woman participant).

Place of origin. Adult women using the SRH services felt that discrimination and abuse they experienced were not related to their Venezuelan nationality. Instead, they believed that mistreatment was a general issue affecting all women. One adult woman shared her experience, “The nurses told me, get up, get up, hurry up, hurry up, we have to take off all those sheets and change them. They treated all badly all of us women who were recovering after our birth; it wasn’t because we were Venezuelan” (Adult women).

Age: Age was another factor influencing perceptions of discrimination and mistreatment. For instance, teenage women reported feeling particularly vulnerable and unsupported, especially in postnatal care settings. They felt that their youth and inexperience were not taken into consideration, leading to harsher treatment from some health professionals.

#### Recommendations to improve access to SRH and reduce discrimination

To enhance access to SRH services for the migrant population in Quito and eliminate discrimination and mistreatment, participants identified several key areas for intervention. These areas included training, education, policies and strategies for comprehensive, inclusive and specialized care, and streamlining access to health services. The following are proposed strategies to improve SRH service access and reduce discrimination:

- Implement comprehensive sexual education in the educational curriculum at all levels, targeting students, parents and teachers to foster better understanding and acceptance of SRH issues (Suggested by Adolescent men – LGBTIQ+ population).
- Provide training for healthcare personnel to ensure healthcare providers offer respectful and effective care and build positive relationships with users (Suggested by Adolescent women).
- Health professionals should provide clear information including details about the access process and the specific interventions that will be performed to help patients understand and feel more comfortable (Suggested by Adult women).
- Healthcare institutions should better understand the needs of the population and allocate a larger budget accordingly to ensure services are adequately funded and can meet demand (Suggested by Adolescent men).
- Healthcare institutions should develop interventions that specifically address the needs of adolescents to prevent early pregnancies (Suggested by Adolescent men).
- Health services should offer friendly, compassionate, equitable care characterized by effective communication and without discrimination, making them more accessible (Suggested by Adult men and Adolescent women).
- The Ministry of Health should prioritize free comprehensive and specialized healthcare for the population of sexual and gender diversities to ensure they receive the care they need without discrimination (Suggested by LGBTIQ+ Population).
- Improve the appointment scheduling system to ensure that people can easily schedule and attend their appointments (Suggested by Adult men).

## DISCUSSION

This study aimed to explore the barriers Venezuelan migrants face in accessing SRH services along their migration trajectory, with a focus on those residing in Quito. Grounded in the perspectives of adult women and men, adolescent girls and boys, and members of the LGBTIQ+ community, this research offers important insights into the complex intersection of sociocultural, economic, ethnic, gender, age-related factors that shape access to SRH service and raises important questions for future inquiry.

### Socio-cultural and economic barriers

The study reveals the socio-cultural barriers such as social taboos surrounding SRH, significantly hinder access to these services. These barriers are compounded by economic constraints, particularly among migrants, who often lack financial resources to seek private healthcare. This finding aligns with existing literature that underscores the inequalities between migrants and non-migrants in accessing SRH services. The socio-economic challenges are further intensified by gender dynamics, with men often neglecting their health due to hegemonic masculinity norms, and women facing financial and social constraints. [18]

### Impact of the COVID-19 Pandemic

The COVID-19 pandemic exposed and exacerbated longstanding deficiencies in health systems across Latin America, including gaps in infrastructure, supplies, workforce capacity, and medical service delivery [19,20]. In Ecuador, already overstretched health services were further strained, leading to deprioritization of many essential services, including SRH care [21,22]. Migrant populations were disproportionately affected, as the crisis compounded their pre-existing vulnerabilities and intensified xenophobic and racist attitudes [21].

A major consequence of the pandemic response was the widespread reallocation of resources toward COVID-19 care, often at the expense of SRH services. The failure to universally recognize SRH as essential care meant that migrants were frequently denied services unrelated to COVID-19, even when urgently needed. This disruption deepened existing barriers to SRH and widened health inequities [21]. The findings underscore the urgent need for inclusive, migrant-sensitive health policies that protect access to comprehensive SRH services, especially during public health emergencies.

### Discrimination and Xenophobia

Participants reported experiencing significant discrimination and mistreatment that hindered their access to SRH services. The nature of this discrimination differed by gender identity, place of origin, and age. For example, the LGBTIQ+ participants described bias largely rooted in their gender identity, while women reported mistreatment directed at all female patients, irrespective of their nationality. These accounts underscore how pervasive discrimination restricts SRH service use.

A critical driver of this problem may be the limited training much health personnels receive on caring for marginalized populations [23]. Transgender individuals, in particular, often avoid seeking medical care because they anticipated prejudice; when they do access services, they may feel obligated to educate providers about their sexuality and health needs [24].. This knowledge gap can lead healthcare providers to wrongfully attribute unrelated health issues to an individual’s sexual or gender identity [24]. One transgender participant, for instance, was questioned about HIV during a consultation for stomach pain, an example of how bias distorts clinical judgement.

Social stigmas likewise affect men, frequently delay or forgo care because prevailing masculinity norm discourage help seeking.[25]. This reluctance intersects with ethnicity and migration status, with men often directing their scarce household income toward family needs rather than their own healthcare. Together, these findings highlight the urgent need for provider training in culturally competent, gender-affirming care and for policies that reduce financial and social barriers to SRH services for all migrant populations.

### Obstetric and gender-based violence

Women, particularly those using family planning and obstetric services, reported experiencing gender-based violence in the form of obstetric violence. This includes abusive and dehumanizing treatment and/or excessive medicalization by healthcare professionals, leading to loss of autonomy and negatively impacting the women’s quality of life[26]. Such mistreatments occurred not only during childbirth but also in prenatal and postnatal care. Study participants emphasized that this poor treatment was directed at all women, regardless of nationality.

### Access for young people

Young participants highlighted barriers to accessing family planning services, particularly the reluctance of healthcare providers to offer adequate information and care. This aligns with global evidence showing that a significant proportion of sexually active adolescents lack access to contraceptives and sexual health information [27]. Fewer than 50% of sexually active women between the ages of 15 and 19 have access to contraceptives,[27]and in a study of 70 low-and middle-income countries, only 10% of adolescent women had visited a health service in the previous year and received family planning information (23). The pandemic exacerbated these barriers, and challenges have persisted in the post-pandemic period.

## Conclusions

This study makes a significant contribution to understanding Venezuelan migrants’ perceptions and experiences regarding access to SRH services. It captures their realities across the migrant journey, from the country of origin, through transit, to settlement, offering a comprehensive view of both the barriers and opportunities.

By focusing on the Venezuelan population in Quito, this study addresses a critical information gap in Ecuador, especially regarding access to SRH services during overlapping crisis such as migration and the COVID-19 pandemic. Findings highlighted how economic instability, discriminatory practices, and inadequate health policies intersect to shape access.

The study identifies several priority areas for future action to ensure unrestricted SRH rights for migrant populations. These include the need for adoption of inclusive health policies, strengthening training for healthcare providers, comprehensive sexual education, and the development of strategies to prevent discrimination to improve enhancement access to SRH services. Addressing these systemic issues could foster a more equitable healthcare system that better meets the needs of both the local and migrant populations in Quito, ultimately reducing discrimination and mistreatment.

## CRediT authorship contribution statement

**José Andrés Ocaña Navas:** Methodology, investigation, data curation, formal analysis, writing – original draft and writing-review & editing. **Susana Guijarro:** Project administration, funding acquisition and investigation. **Gabriela León:** Data curation and formal analysis **Ana Lucía Torres:** Investigation, supervision, methodology and project administration. **Patricio Murgueytio**: Project administration, methodology and funding acquisition. **Odalys Heredia-Andino:** writing-review & editing. **Paula Yánez-Sánchez:** writing-review & editing**. Sarah Iribarren:** investigation, supervision, methodology, funding acquisition and writing-review & editing.

## Data Availability

The data supporting the findings of this study cannot be made publicly available due to ethical restrictions. Although the data are anonymized, they consist of sensitive qualitative information concerning sexual and reproductive health, and were collected from a socially vulnerable migrant population, which included minors. However, the data are available upon request for researchers who meet the criteria for access to confidential data. Requests should be directed to the Data Analysis Unit of the Institute of Public Health at the Pontificia Universidad Católica del Ecuador. Please contact and cc the University's Ethics Committee at.

